# Sustained effects of Short-term Adalimumab biosimilar (ZRC-3197) therapy on outcome measures in Ankylosing Spondylitis

**DOI:** 10.1101/2021.11.14.21266244

**Authors:** Chopra Arvind, N Khadke, M Saluja, T Kianifard, Mihir Gharia, A Venugopalan

**Author notes:** Address for Communication: Dr Arvind Chopra, Centre for Rheumatic Diseases., Hermes Elegance, 1988 Convent St., Camp, Pune, 411001 India. Drug Trial Registration: Clinical Trial Registry of India vide reference CTRI/2018/02/012018 and available at http://ctri.nic.in/Clinicaltrials/pmaindet2.php?trialid=19498&EncHid=&userName=adalimumab. **Disclosures:** All the authors of the current study are full time employees of CRD Pune and did not receive any personal payment or compensation from any source towards the current study or current publication. **Funding:** Zydus Cadila India donated a substantial quantity of biosimilar Adalimumab (Exemptia™) injections for free use by the deserving and needy patient participants in the study and further provided an unrestricted educational and research grant to Arthritis Research Care Foundation-Centre for Rheumatic Diseases CRD), Pune, India. **Availability of data and material:** All study related medical and other records are archived in Centre for Rheumatic Diseases (CRD), Pune, India. An e-database was created to capture the data required for study analysis and access will be provided one year after publication after a formal request inclusive of purpose of data access to the first author (Dr Arvind Chopra) of the current study. **Authors contributions:** Conceptualization: Arvind Chopra; Methodology: Arvind Chopra, Manjit Saluja, Nagnath Khadke, Toktam Kianifard, Anuradha Venugopalan; Formal analysis and investigation: Arvind Chopra, Anuradha Venugopalan; Writing - original draft preparation: Arvind Chopra; Writing - review and editing: Manjit Saluja, Anuradha Venugopalan; Funding acquisition: Arvind Chopra; Resources: Arvind Chopra, Anuradha Venugopalan, Manji Saluja: Supervisor. **Ethics approval:** Approval was obtained from the ethics committee of the Centre for Rheumatic Diseases, Pune, India, and the procedures used in this study adhere to the tenets of the Declaration of Helsinki. Arvind Chopra, Manjit Saluja. **Consent to participate:** Informed consent was obtained from all individual participants included in the study. **Consent for publication:** No specific data or photograph of any kind of a study participant is used in this study.

## Abstract

**Introduction:** Cost and drug toxicity often deter prolonged therapeutic use of anti-TNF agents in ankylosing spondylitis (AS). A planned study was completed to endorse our clinic-based observation of long-term relief following short-term administration of an anti-TNF agent.

**Methods:** 50 consenting patients with symptomatic active chronic AS under rheumatology care in a community clinic were enrolled; naïve for anti-TNF. 40 mg standard biosimilar Adalimumab (Bs-ADA, Exemptia™) was injected subcutaneously every fortnight for six injections (10 weeks). Patients were monitored at several predetermined time points. Improvement was assessed with standard indices (Assessment Spondyloarthritis International Society/ASAS and Bath). An intention to treat analysis was performed: significant p <0·05

**Results:** Patients showed early and substantial significant improvement in pain, NSAID requirement, function, and in several indices (ASAS 20 & 40, ASAS partial remission, BASDAI, BASFI, ASDAS) which persisted after stopping injections. 84% and 52 % of patients respectively showed ASAS 20 improvement at weeks 12 and 48: corresponding to ASAS partial remission at 34% and 24%. Over 50% of patients maintained prolonged improvement and provided proof of concept (defined apriori). Serum Interleukin-6 assay showed a sharp reduction at 24 weeks. None developed TB or serious drug toxicity. 11 patients withdrew (mostly inadequate response). The absence of control was a limitation.

**Conclusion:** A ten-week administration of biosimilar adalimumab in difficult-to-treat AS showed early substantial improvement which often persisted for 24 weeks. This unconventional strategy was socioeconomically appealing. It merits further validation and acceptance, especially in resource strained settings.

## Introduction

Ankylosing spondylitis (AS) is a prototypic autoimmune inflammatory painful disorder of the axial skeleton and sacroiliac joints and causes substantial limitation in spine motion, spine deformity, systemic complications, and premature mortality [1]. The newer imaging modalities offer early diagnosis and assessment of skeletal damage [2]. AS is a lifelong and difficult-to-treat disorder [3]. NSAID (non-steroidal anti-inflammatory drugs) remains the mainstay of symptomatic treatment. The benefit of conventional DMARD (Disease-modifying anti-rheumatic drugs) is debatable, whereas anti-tumor necrosis factor (TNF) drugs and other biological DMARD are more effective and recommended for long-term use [3]

The prevalence of AS in India was reported to be 0·03% (95% confidence interval 0·02, 0·05) but this would mean a substantial disease burden [4]. The unaffordability and risk of infections including tuberculosis (TB) remain a deterrent to the widespread use of biologic DMARDs in India.

Several biosimilar anti-TNF drugs were launched globally to improve the access and affordability (5, 6). Adalimumab (Humira™) is a popular anti-TNF drug but not marketed in India. In 2014, a standard biosimilar Adalimumab (Exemptia™, Bs-ADA) was marketed in India [7,8] A generous donation of Exemptia™ was instrumental in facilitating the current non-commercial investigator-initiated study.

A repeatedly observed prolonged symptomatic relief following a shorter period (non-standard) of treatment with Infliximab or Etanercept in several patients of AS in clinical practice inspired the author (AC) to experiment with a new concept. A preliminary report was presented at the 2005 APLAR Congress [9]. Short-term therapy seemed effective and safe, and certainly more acceptable to the patients. The current study was planned with this perspective in mind.

The aim of the study was to evaluate the prolonged effectiveness and safety of a short-term regimen of a biosimilar Adalimumab in the treatment of active chronic symptomatic AS.

## Patients and Methods

This was a prospective interventional uncontrolled single centre study of one year duration. The design was observational and it was carried out in a popular community rheumatology clinic (Centre for Rheumatic Diseases, Pune, India) from June 2016 to July 2018. The protocol adhered to the guidelines of the updated Declaration of Helsinki and was approved by the local ethics (CRD, Pune) committee and registered (CTRI/2018/02/012018) [10].

Inclusion criteria included (i) clinical diagnosis of AS made by a rheumatologist as per ASAS criteria (ii) Patients aged between 18 to 75 years (iii) symptomatic disease and fulfilling any two of the following three criteria anytime during the previous week (a)muscular-skeletal axial pain VAS of 3 cm or more (range 0 to 10 cm) for over three months (b) early morning stiffness of 30 min or more (c) Erythrocyte Sedimentation Rate (ESR, Westergren method) ≥ 28 mm end first hour or C-Reactive protein assay (CRP) ≥ 6 mg/l and (iv) bony erosions and or ankylosis as evident from a plain digital standard skiagram of the pelvis for sacroiliac joints (v) patients requiring frequent NSAIDs who have been on a stable dose for at least 4 weeks (vi) duration of disease ≥ 2 years.

Exclusion criteria included (i) arthritis and/or axial spine pains due to any other cause except reactive arthritis (ii) biological agent use (especially anti-TNF) during the previous two years (iii) DMARD use except sulphasalazine and or methotrexate during the previous 12 weeks (iv) steroid use in the previous 4 weeks (v) recent onset of a potentially serious medical disorder (such as diabetes or cardiovascular disease) (vi) history of any recent infection requiring antibiotics in the last 12 weeks (vii) history of past tuberculosis irrespective of the nature of treatment provided (viii) past history of hepatitis or human immunodeficiency virus (HIV) (ix) history of cancer or lymphoproliferative disease (x) exposure to any kind of hazard or risk by virtue of participation in the study as judged by the investigator

### Selection, Screening, and Enrolment

Potential patients were identified from the outpatient clinic (CRD, Pune) where a comprehensive clinical database exists since 1998. After signing the informed consent form patients were examined for eligibility and enrolled on a first-come-first-serve basis.

### Study Visits

After enrolment, patients visited the study site (CRD) every two weeks during the initial 10 weeks to receive the Bs-ADA injection (total 6 injections) and subsequently at 12, 24, 36, and 48 weeks (one-year study completion) endpoint.

### Study and Concomitant Medication

40 mg Bs-ADA (Biosimilar Adalimumab) was injected subcutaneously every fortnight as per protocol. Patients who failed to achieve ASAS 20 index response at week 12 were randomized to receive two more fortnightly injections (Bs-ADA) or continue routine follow-up. BS-ADA was not repeated ever thereafter.

Patients were counseled on pain relief and the use of NSAIDs (AC, NK). They have prescribed a daily therapeutic dose of NSAID for the initial 4 weeks and thereafter switched to a need-for basis (prn) as judged by the rheumatologist (Table 2). Patients were encouraged to record daily NSAID use in a personal diary. Patients were allowed oral paracetamol (one 650 mg tablet 1-2 times daily) and or tramadol (50 mg tablet, 1-2 times daily) for intolerable pain. Patients were monitored every two weeks for the initial twelve-week period.

Most of the patients used Etoricoxib (60 mg and 90 mg tablets, India Pharmacopeia) prior to enrolment and were permitted to continue the same (initially 60-120 mg daily) after enrolment (Table 2). The use of other NSAIDs was permitted but the dose was converted into Etoricoxib equivalent for the purpose of compiling the mean daily NSAID dose data (Table 2). One thousand milligrams of Naproxen or 150 mg of Diclofenac or 100 mg of Indomethacin or 900 mg of Etodolac was considered equivalent to 120 mg of Etoricoxib as decided apriori by author (AC).

Steroids in any form were not permitted. Patients continued medicines for any other coexistent disease with primary care physician.

### Efficacy Measures

A routine standard rheumatology case record form and another standard AS questionnaire, including those recommended by ASAS (Assessment Spondyloarthritis International Society) were employed [2, 11-14]. Patients also completed the Indian version of HAQ (modified Stanford Health Assessment Questionnaire) [15].

Important measures were as follows: (i) VAS (visual analog scale) was used to record musculoskeletal pain (maximum pain experienced in the previous 24 hours), patient and physician global assessment (current) and general health (current). It was a horizontal line anchored at 0 for nil and 10 for optimum value (ii) NSR (numeric scale rating) with 10 marked categories (1-10) on a horizontal scale (iii) BASDAI (Bath disease activity index): 6 questions marked on NSR to evaluate symptoms pertaining to previous week - fatigue, neck-back-hip pains, pain/swelling in peripheral joints, discomfort on eliciting local tenderness (entheses), duration and severity of early morning stiffness (iv) BASFI (Bath functional index):10 items marked on NSR for daily bodily activities such as climbing steps, doing work related tasks etc (v) ASAS 20 index improvement response was the primary efficacy variable and evaluated 4 domains-patient global assess, overall pain, function (BASFI) and inflammation (mean of BASDAI questions 5 & 6 pertaining to morning stiffness); achieved with a minimal 20% improvement with at least 1 unit improvement in NSR in 3 of the 4 domains without any worsening in the remaining domain (20% or more and more than 1 unit) (vi) ASAS 40 index improvement response: domains similar to ASAS 20 but requiring 40% improvement and minimal 2 unit improvement in NSR and without any worsening (vii) ASAS partial remission required none of the 4 domains in ASAS 20 to exceed 2 on the NSR (viii) ASAS 5/6 response required at least 20% improvement in 5 of the 6 domains (ASAS 20 domains plus domains of lateral spine flexion and CRP assay) (ix) ASDAS (disease activity score), a weighted index, was computed using a standard online program and included total back pain, patient global disease activity, peripheral painful/swollen joint, morning stiffness and ESR.

Patients were required to report to the study site in a fasting state (without the intake of a meal or any study drug) during the morning hours (8-11 am) of the scheduled visit.

Safety Evaluation: Adverse events (AE) were recorded in the CRF

### Laboratory Evaluation

Patients completed laboratory investigations during screening and at baseline and at week 4, 8, 12, 24, 36, and 48 (one year) endpoints as per protocol. These included routine hemogram, blood sugar, metabolic hepatic and renal profile, urine analysis, Erythrocyte Sedimentation Rate (ESR, Westergren method, normal range: for men-less than 20 mm and for women-less than 30 fall end of 1^st^ hour) and C - reactive protein titter (CRP, Nephelometry, normal range 0-5 mg/dl). Patients were screened for latent TB using standard skin tuberculin testing (5 TU intradermal), blood TB (gamma interferon release assay and skiagram of the chest [16]. A commercially available ELISA kit was used to assay selected cytokines-TNF alpha, Interleukin (IL)-6, and IL-17.

### Patient Expenditure

The Biosimilar Adalimumab (Exemptia™) was provided free of cost to 13 patients and at almost 50% concessional market rate to the rest of the participants in the study. Patients purchased their own NSAID and analgesic. All investigations were carried out free of cost. Patients were given a modest monetary allowance for travel and meals.

### Statistical design and data analysis

A convenient (non-probabilistic) sample size was chosen by AC (lead author) based on local resources but with an emphasis on at least 35 participants. A 20% dropout rate was anticipated by AC based on earlier experience. Appropriate statistical tests were performed using a standard statistical package (SPSS version 2007, USA) with significance at p< 0.05, two-sided. Quantitative variables were analyzed for mean, median, standard deviation (SD), proportions, and 95% Confidence Interval (CI). An intention to treat analysis (last observation carried forward) was completed. ASAS 20 was the primary efficacy response at week 12, 24, 3,6 and 48 endpoints The proof of the concept (see above) in the current study was to be accepted if at least 50% of the patients with ASAS 20 response at week 12 (two weeks after cessation of treatment with Bs-ADA) showed persistence of response for another 24 weeks (at week 24 and week 36 endpoints).

A conventional survival analysis (life table method) was also carried out with the ‘survival’ defined as an absence of the event (such as ASAS 20, ASAS 40, ASAS 5/6 response, ASAS partial response) that persisted for at least eight weeks.

### Observation and Results

Fig 1 shows the disposition of patients in the study. 60 patients were screened and 50 patients (42 men, 8 female) were found eligible and enrolled. In retrospect, all patients satisfied the standard classification criteria for AS [2, 17]. The baseline characteristics of all patients are presented in Table 1.

**Table 1:** Demographic Data

| BASELINE CHARACTERISTICS | AS (n=50) |
| --- | --- |
| Age (in years) (Mean±SD) | 31.2 (3.53) |
| Gender (M/F) | 42 / 8 |
| HLA B27 Positive (%) | 86%(n=43) |
| Body weight (Kg) (Mean+SD) | 63.1(14.5) |
| ESR mm/hr (Mean+SD) | 88±30 |
| CRP mg/L (Mean+SD) | 63.8 ± 55.2 |
| Duration of Disease(years) | 31.2 (SD ±9) |
| On DMARDs | 28 |
| On NSAIDs | 9 |
| VAS (Pain) (Mean+SD) | 5.9±2.0 |
| BASDAI (Mean) | 5.3 |
| BASFI (Mean) | 4.7 |
| PATG (Mean+SD) | 6.2±1.6 |
| HAQ (Mean+SD) | 5.3±3.7 |

**Fig 1:**
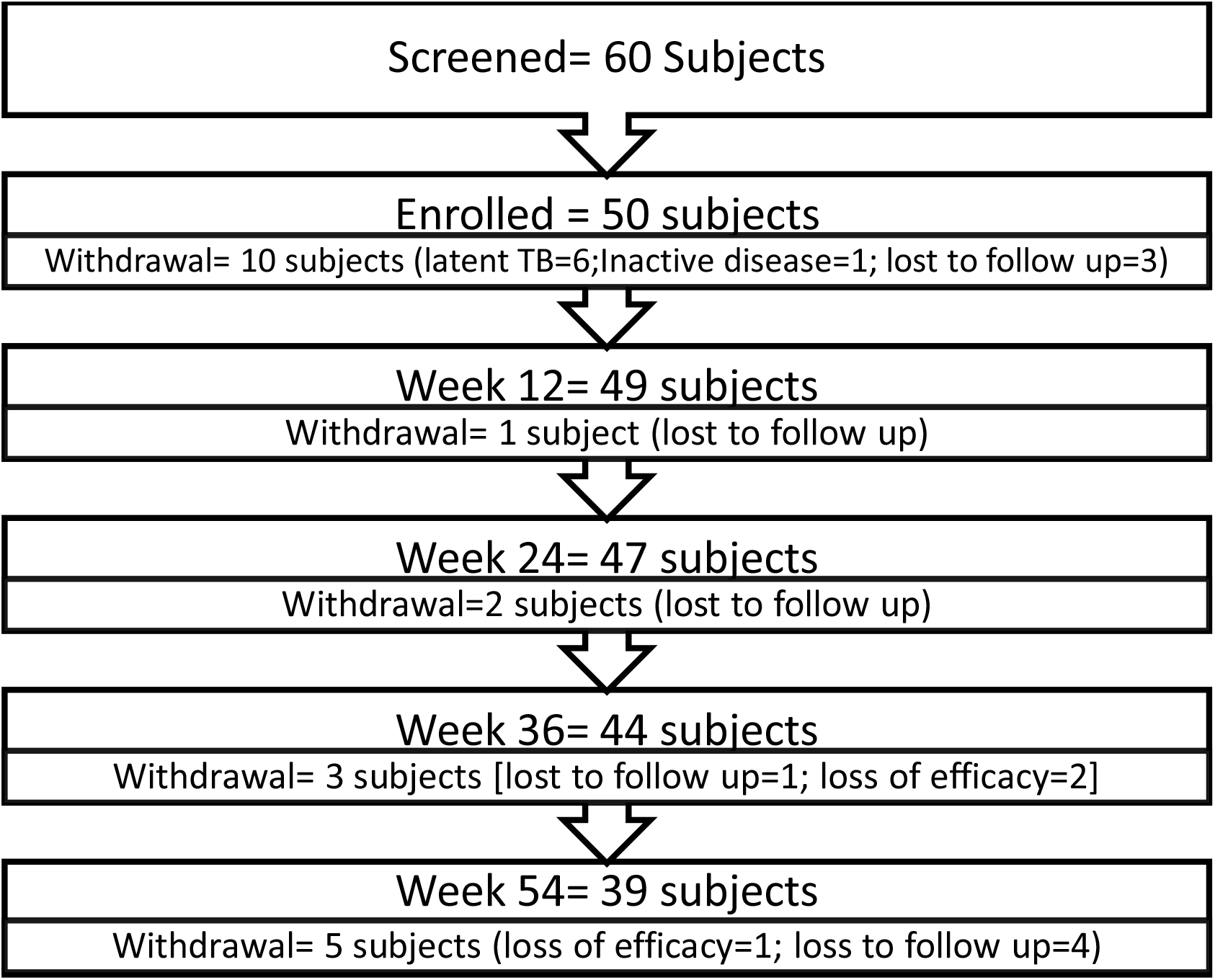
Patient disposition flow chart: A study of prolonged benefit of a short term administration of biosimilar Adalimumab in active symptomatic Ankylosing Spondylitis [Adalimumab administered for initial 12 weeks in 46 patients, 4 weeks in 1 patient, 16 weeks in 3 patients, see text for details].

The mean age and duration of illness were 31·2 (SD ±9, Range 19-56) years and 99·5 (SD± 67.8, Range 13-312) months, respectively. 86% of patients tested positive for HLA B27. All patients had consumed multiple courses of more than two NSAIDs, often supervised, during the recent past period as per medical records. The NSAID use and consumption are shown in Table 2. 28 patients provided a past history of a supervised prolonged (at least six months) intake of standard DMARD (sulphasalazine and/or methotrexate).

**Table 2:** NSAID (Etorocoxib or Etoricoxib equivalent) consumption during the trial study of biosimilar Adalimumab (Bs-ADA) in patients (n=50) suffering from symptomatic moderately severe chronic Ankylosing Spondylitis: A single-arm prospective observational study

| Weeks, 0 Baseline / Variables | 0 to -4* | 0 to 2 | 2 -4 | 4-6 | 6-8 | 8 -12 | 12-24 | 24-36 | 36-48 | 48+ |
| --- | --- | --- | --- | --- | --- | --- | --- | --- | --- | --- |
| Number patients | 50 | 50 | 50 | 49 | 49 | 49 | 49 | 47 | 44 | 39 |
| Mean daily dose mg (SD) | 92.4 (27.62) | 104.7 (25.3) | 108.7 (31) | 77.3 (46.7) | 35.1 (42.4) | 19.6 (35.7) | 17.2 (32,2) | 22.2 (38.9) | 26.6 (42.8) | 50.5 (50.1) |
| **Continuous use | 50 | 50 | 45 | 37 | 17 | 9 | 4 | 6 | 9 | 17 |
| ***On need basis | 0 | 0 | 5 | 7 | 13 | 15 | 14 | 15 | 19 | 11 |
| ^Nil use | 0 | 0 | 0 | 5 | 19 | 25 | 31 | 26 | 16 | 11 |

Note: Bs-ADA was injected using a standard regimen for 12 weeks beginning at O Baseline ;NSAID: non-steroidal anti-inflammatory drug; Mean daily dose pertains to Etorocoxib (Nucoxia™/ Ezycoxib™ & available as 60 mg and 90 mg tablet, India pharmacopeia); Over 90% patients consumed Etorocoxib in the study; Dose of any other NSAID converted empirically to Etorocoxib equivalent ;\*: minus prefix indicates period prior to baseline; 48+; study completion and a 12 week follow up; \*\*: either od or bid, 60-120 mg Etoricoxib;\*\*\*: 60 mg Etorocoxib od on need basis; ^ :Advised need basis but did not require any NSAID use; See text for details especially for Etoricoxib equivalent

Patients showed a moderately severe symptomatic active disease as shown by pain VAS and other individual variables (Table 3). The baseline mean spine pain, based on the BASDAI questionnaire, was 6.8 ±2.1 (SD) (range 0-10 cm). 42 patients (84%) were classified with a ‘very high disease activity state as per ASDAS [14].

**Table 3:** Efficacy variables (Mean ± Standard Deviation) at baseline and predetermined study time points in a study of biosimilar Adalimumab (administered for initial 12 weeks) in Symptomatic Ankylosing Spondylitis (n= 50 patients) - a single arm prospective observation study (95% confidence interval shown in parenthesis)

| Variable/<br>Time<br>points | Baseline | Week 4 * | Week 8* | Week12 * | Week 24 | Week 36 | Week 48 * |
| --- | --- | --- | --- | --- | --- | --- | --- |
| Pain<br>VAS | 5.9±2.0<br>(5.3, 6.5) | 3.7±1.7<br>(3.3, 4.2) | 3.2 ±1.9<br>(2.6,3.7) | 2.6±2.1<br>(2,3.2) | 3±2.4<br>(2.3,3.7) | 3.6±2.4<br>(2.9,4.2) | 3.92±2<br>(3.3,4.6) |
| PATG | 6.2±1.6<br>(5.7, 6.6) | 4 ±1.5<br>(3.6, 4.5) | 3.3±1.6<br>(2.9,3.8) | 2.9±1.9(2.3,<br>3.4) | 32(2.6, 3.5) | 3.5±2.1<br>(2.8, 3.9) | 3.72±3<br>(3.1,4.4) |
| HAQ | 5.3±3.7<br>(4.2, 6.3) | 3.1±2.8<br>(2.3, 3.9) | 2.5±2.4<br>(1.8,3.1) | 2.1±2.8<br>(1.4,3) | 3<br>(2.2,3.9) | 3.5±3.2<br>(2.6,4.4) | 3.4±4<br>(2.5,4.4) |
| ESR | 88±30<br>(80, 97) | 29.1±20<br>(23.4, 34.8) | 31.4±20<br>(23.4,34.8) | 33.929.1<br>(25.6,42.1) | 54±33.2<br>(43.4,62.3) | 69.837.2<br>(59.3,80.5) | 68.5±33.2<br>(59,78) |
| CRP | 63.8 ± 55.2<br>(48.1, 79.5) | 9.6 ± 16.8<br>(4.8, 14.4) | 13.2 ± 29.6<br>(4.8, 21.6) | 10.9 ± 21.4<br>(4.8, 17.0) | 25.0 ±39.4<br>(13.8, 36.2) | 35.0 ±44.8<br>(22.3, 47.8) | 35.6 ± 38.0<br>(24.8, 46.4) |

Note: See text for methods; All numerical values rounded to 1 decimal place; n: number; \*: significant difference from baseline at $p < 0.05$ (ANOVA), intention-to-treat analysis; VAS: visual analogue scale; BASDAI: Bath disease activity index; ASDAS: Ankylosing Spondylitis disease activity score; PATG: patient global assessment; HAQ: Health assessment questionnaire (Indian version); ESR: Erythrocyte sedimentation rate (Westergren, mm fall end 1<sup>st</sup> hour);CRP: C-reactive protein (mg/l); Hb: haemoglobin (mg/dl)

### Withdrawals

11 patients withdrew and mostly because of poor therapeutic response. None of the withdrawals were due to an AE (Fig 1). 1 patient developed intolerable hip pain and underwent hip arthroplasty after 30 weeks of study.

### NSAID

Table 2 shows the quantitative use of NSAID (Etoricoxib). 9 patients used other NSAIDs (3 Indomethacin, 3 Naproxen, 2 Diclofenac sodium, 1 Etodolac) and their dose was converted to Etoricoxib equivalent (see method). The mean pain VAS after two weeks was 4.2±2 (SD). However, a substantial reduction in the use of NSAID was observed after patients completed 4 weeks of study (3 Bs-ADA injections). Less than 15% of patients required daily NSAID after 8 weeks. 4 of the nine patients who consumed NSAID daily at week 12 showed ASAS 20 index response.

### Effectiveness (Table 3, Table 4)

The ASAS 20 improvement at week 12 was seen in 41 (82%) patients (95% CI 69.2%, 90%) and ASAS 40 in 35 (70%) patients (95% CI 56.2 %, 80.9%). Over 50% of patients achieved ASAS 20 response at all primary efficacy time points (Table 4). 21 patients (42%) showed persistence of ASAS 20 index improvement at week 12-, 24-, and weeks time points and provided proof of the concept (see the statistical design and data analysis).

**Table 4:**
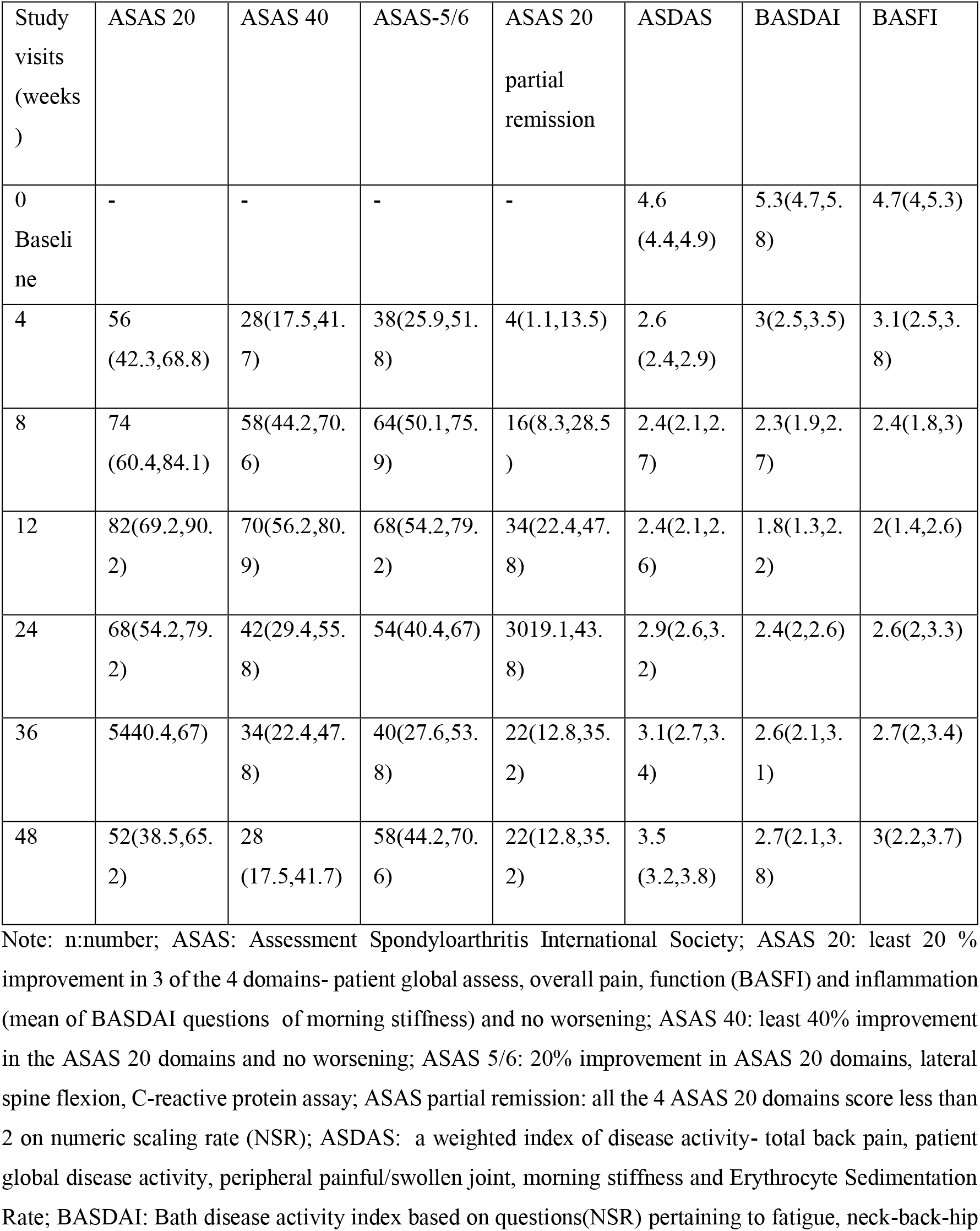
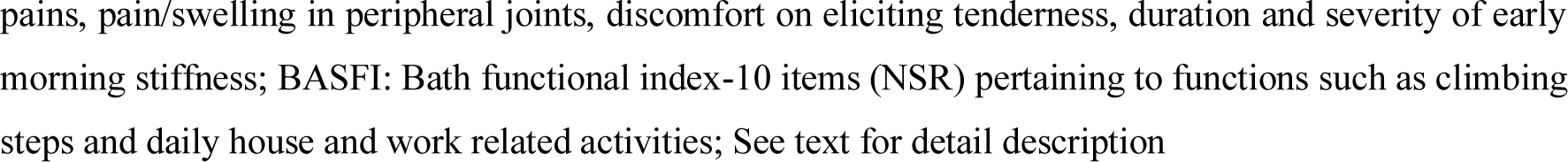
Proportion percentage (95% confidence intervals) of patients showing ASAS 20 (Primary efficacy response), ASAS 40, ASAS 5/6, and ASAS partial remission and mean of ASDAS (ESR), BASDAI and BASFI indices at predetermined study time points in a study of biosimilar Adalimumab (administered during the initial 12 weeks of study) in Ankylosing Spondylitis (n= 50 patients)-a single arm prospective observational study (95% Confidence interval shown in parenthesis, intention to treat analysis)

Patients improved significantly with the early response which persisted, albeit with some decline, till the study completion at 48 weeks (one year) (Table 3, Table 4, Fig 2) the optimum improvement was shown at week 12. Based on ASDAS, inactive/low disease activity was classified in 28 patients (56%) at week 12, 18 patients (36%) at week 24, 12 patients (24 %) at week 36, and 10 patients (20 %) at week 48 endpoint; corresponding ‘major improvement’ seen in 64%, 50%, 36%, and 28% patients [14].

**Figure 2:**
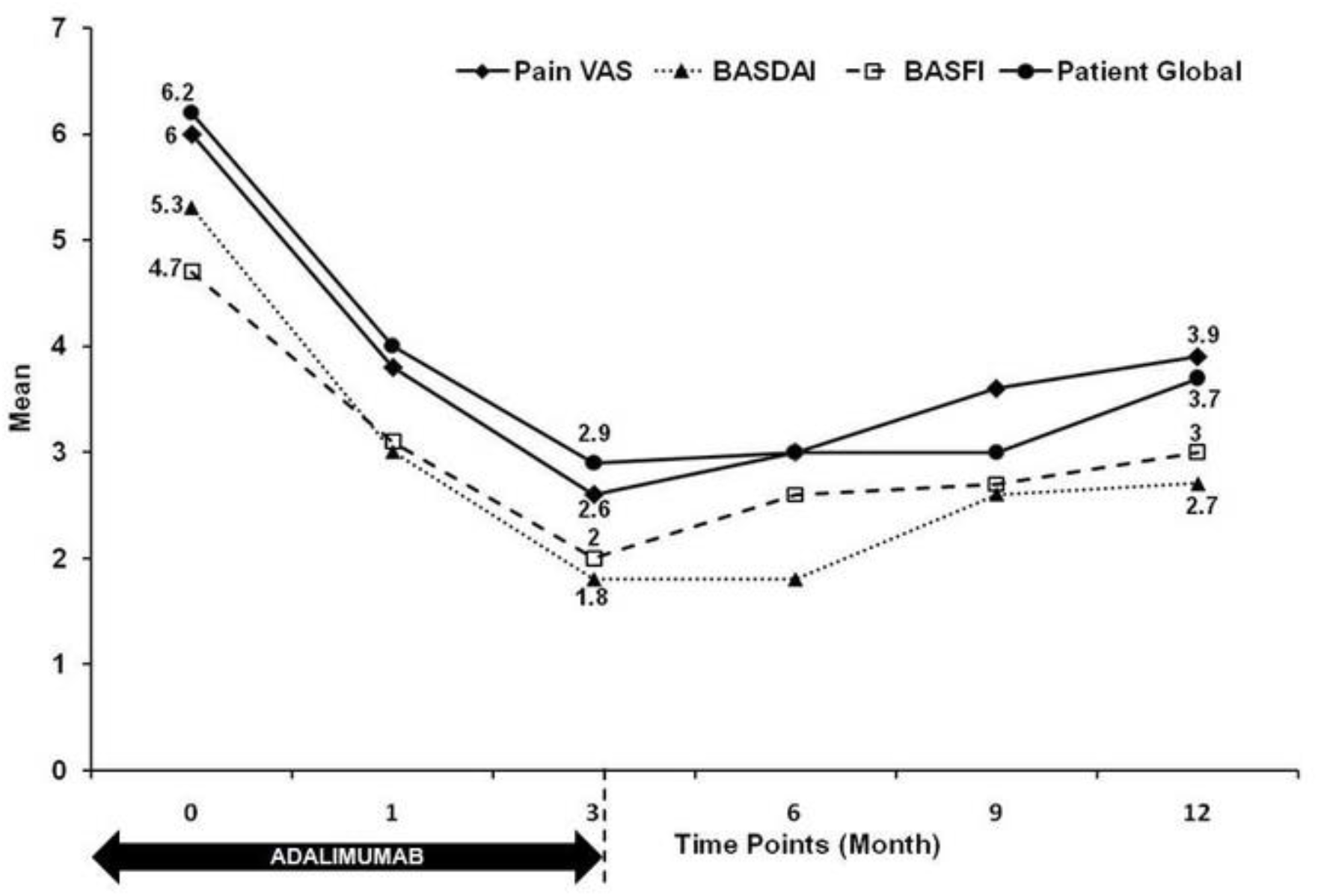
Selected Efficacy Measures (Mean) over time: An open label study of biosimilar Adalimumab in Ankylosing Spondylitis (n= 50 patients)[n:number; VAS: visual analogue scale; BASDAI and BASFI are indices of disease activity and functional impairment; See text for methods].

### Survival Analysis (Fig 3)

ASAS 20 achieved a high cumulative proportion (¬80%) in the cohort within 4-12 weeks of study. ASAS 40 and ASAS 5/6 showed a similar pattern (time to event) but the cumulative proportion was less (¬50%) and the curve remained nearly flat (minimal increment) thereafter. The time to event for optimum (¬30%) cumulative proportion for ASAS partial remission event was seen later at 12-24 week intervals and it remained near flat thereafter (week 36: 95% CI 20%, 45%).

**Fig 3:**
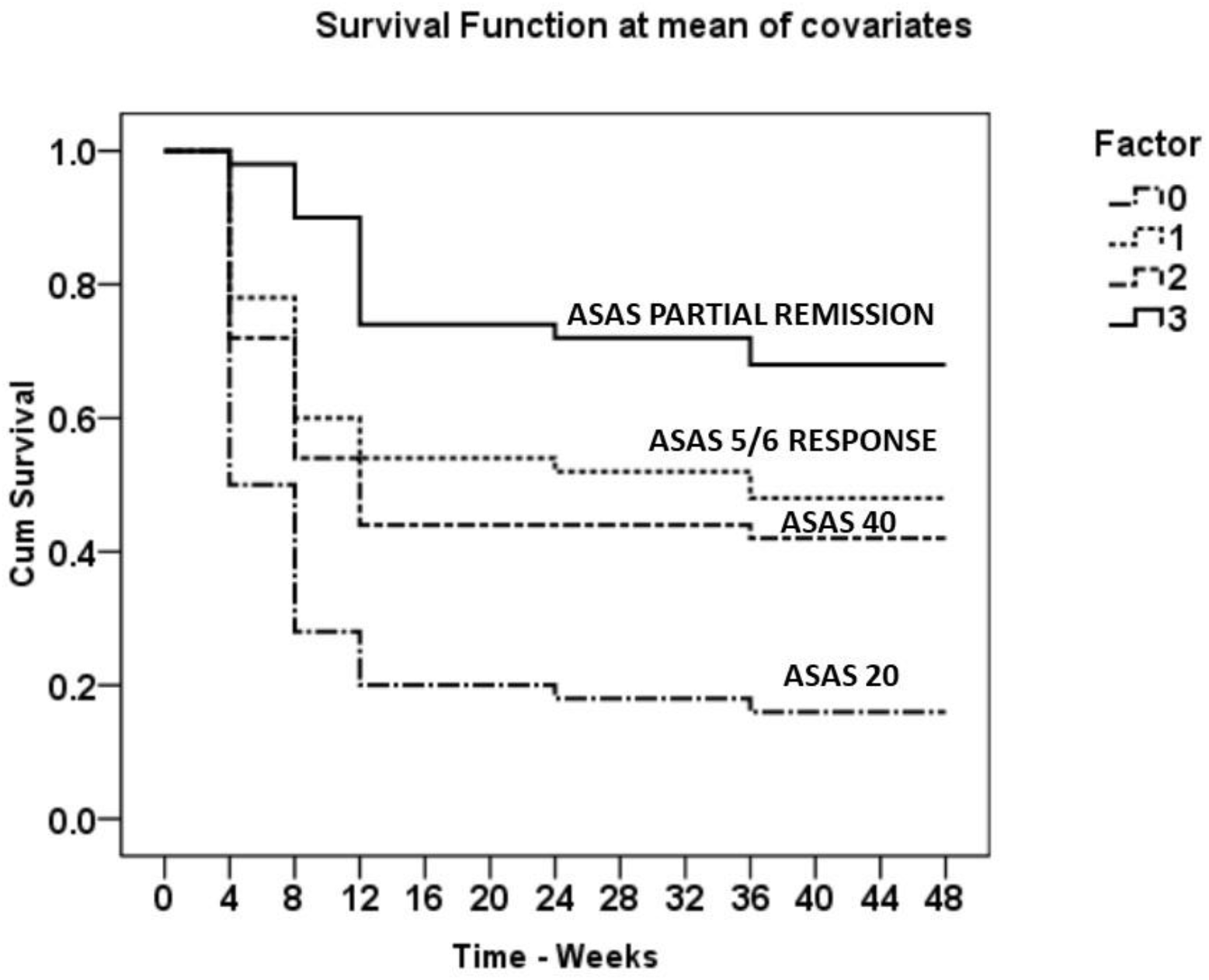
Survival Analysis.

### Non-respondents

Seven patients who were examined at week 12 failed to achieve ASAS 20 response. Despite 3 randomized patients receiving additional two Bs-ADA injections (see methods), none of the non-respondents achieved an ASAS 20 response thereafter.

### Safety and tolerability

None of the patients suffered from serious AE. There were no deaths. None developed symptomatic TB. 10 patients reported mild AE which resolved with standard care and none showed local site reaction (3 patients had nasopharyngitis, 3 patients had dermatophytosis, 2 patients had non-specific body aches and pains, 2 patients had mild uveitis, 1 patient had nasal polyp bleed, 1 patient itchy eczema over ankle region).

### Cytokine Assay (Figure 4)

Pro-inflammatory cytokines assay (IL-6, TNF α, and IL-17) showed a reduction at week 24 and week 48 as compared to baseline and was substantial for IL-6 assay at week 24 (p<0.05).

**Figure 4:**
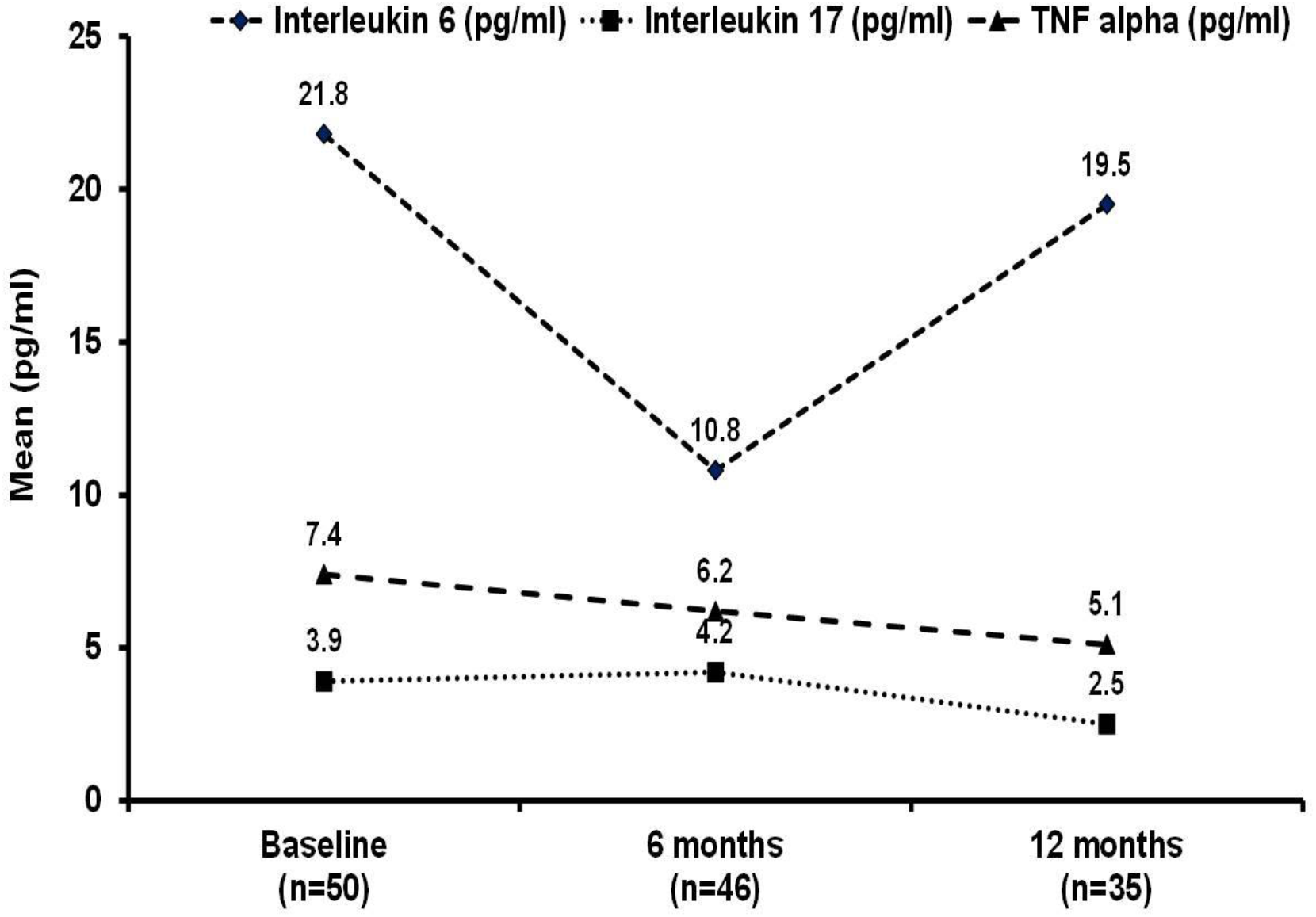
Selected Cytokine Assay (Mean) over time: An open-label study of biosimilar Adalimumab in Ankylosing Spondylitis (n= 50 patients)[n: number; See text for methods)

## Discussion

This prospective observational study of one-year duration demonstrated an early onset of significant and substantial improvement in patients suffering from active chronic symptomatic AS after treatment with a biosimilar Adalimumab administered over 10 weeks (six injections). The patients had shown an inadequate response to NSAID prior to enrolment and the consumption (NSAID) was reduced considerably and persisted over a prolonged period (Table 2). Significant and considerable improvement persisted for several variables and indices (Table 3 and Table 4), albeit with a modest decline after stopping Adalimumab. Eighty-two percent of patients showed ASAS 20 index improvement (primary efficacy) at week 12 and the response persisted in over 50% of patients for at least 24 weeks (proof of the a-priori study concept). 11 patients withdrew because of inadequate improvement. All AE were mild. The study was carried out in a real-life community practice setting.

The extent of improvement desired by a patient suffering from chronic AS is difficult to assess. Several standard index responses (ASAS, BASDAI, BASFI, and ASDAS) were used in the current study to show a multifaceted improvement [2]. These indices also demonstrated persistence of good improvement during the study period as shown by the survival curves (Supplementary Fig 1). There was comparatively less improvement with ASDAS. (Table 4). In our experience, patients often struggle to quantify morning stiffness and we believe that some of the study patients failed to donate blood samples in a fasting state because of difficult personal logistics.

Interestingly, the ASAS partial remission in the current study was 34% at week 12 (Table 4) and more or less persisted after stopping Adalimumab (Supplementary Table 2 and Fig 1). It is difficult to define remission in chronic AS [18]. Up to a third of patients treated with NSAID and up to two-thirds treated with anti-TNF agents are reported to achieve ASAS partial remission with higher responses seen in patients with the more recent disease [18]. Spontaneous remission in AS is rare and partial remission (ASAS) was reported in 1.03-5.6% of patients on placebo in anti-TNF studies [18].

Current recommendations advocate long term use of anti-TNF agents in AS. Several patients relapsed within 6 months of stopping infliximab after using it regularly for three years [19]. But in the current study, several patients continued to enjoy symptomatic improvement (Table 3, Table 4, and Fig 2) with good function (low BASFI) for at least 24 weeks after receiving 6 injections of Bs-ADA. Perhaps short-term therapy (anti-TNF) can be used further in a cyclical manner (10-week therapy every 24 weeks) in some patients with difficult AS resulting in good efficacy and safety. But this needs to be validated in suitable long-term clinical studies.

There were several other limitations and strengths. The study was uncontrolled and carried out in a single centre with a small sample size. The basic concept in the study was inspired by a therapeutic need and the clinical observations of the author (AC) [5]. However, the protocol was pragmatic in nature and patients were managed in a community setting and showed good compliance; 6 of the 11 withdrawals were after 36 weeks of study (Fig 1). Several of the current selection criteria and methods were influenced by the experience of the author (AC) and differed from standard recommendations (ASAS) and regulatory drug trial studies [3, 20]. The placebo effect arising from the patient’s expectations regarding Bs-ADA must have influenced the overall outcome, especially in the initial phase. The current study did not assess anti-ADA antibodies and radiological (structural) changes and the benefit of long-term use of anti-TNF agents on the bony lesions in AS seems to be modest [21]. The current study adhered to the requirements of longitudinal observational drug studies in rheumatology as advocated by EULAR (22).

Prolonged use of NSAID in AS is a standard of care and a good symptomatic response is expected within 2 weeks [3]. A randomized controlled study of a daily fixed dose of Etoricoxib over a one-year duration demonstrated excellent efficacy and safety in patients with AS although the disease duration was not described and both 90 mg and 120 mg groups showed almost a similar outcome [23]. Before enrolment, 77.4% of patients flared considerably following the washout phase. The reduction in pain after two weeks was substantial and persisted till study completion (one year). At week 6, the ASAS 20 index improvement was 64.8% and the ASAS partial remission was about 15%. In another randomized controlled study of 24-week duration, a combination of infliximab and naproxen was significantly superior to naproxen alone and both groups showed significant early pain relief by two weeks and a persistent reduction in BASDAI and CRP; the disease duration was less than 3 years [24].

In the current cohort, patients suffered from active chronic AS and the response to NSAIDs had been inadequate. Etoricoxib is the preferred NSAID in our clinic for AS treatment. Despite previous regular daily use, the baseline (week 0) pain was moderately severe (Table 3). However, patients continued Etoricoxib with the therapeutic dose along with Bs-ADA. The reduction in pain after 4 weeks (-2.2. 95% CI -2.77,-1.65) was considerable and significant (Table 3) and a substantial and persistent reduction in the requirement of NSAID was observed (Table 2). Patients stopped Bs-ADA after week 10. Less than 15 patients admittedly consumed oral paracetamol and/or tramadol infrequently on a need-for basis during the initial 12 weeks (data not shown). Though the selection criteria and study design differed considerably, the extent of active painful disease in the two NSAID drug trials described above was comparable to the current study.

There was no washout period in the current study. The pain relief to NSAID (Etoricoxib) in the current study was relatively less. It is difficult to vouch for the data on NSAID use shown in Table 2 though we are largely certain. It was largely based on patient compliance and recall which can be difficult to judge. The data on the daily use of NSAIDs and absent use is likely more reliable (Table 2). Etoricoxib may not be the best NSAID for all patients. We often encounter patients (and the community) who express intense fear of consuming NSAIDs and analgesics because of potentially life-threatening side effects and so our expert counseling often fails. Yet there is still, a large majority of deserving patients of AS who cannot afford biological agents although they would wish to make use of them. The results of the current study also ought to be considered against this community perspective.

In a randomized active-controlled drug trial study, the ASAS 20 index response at week 24 was 75% of patients in the Biosimilar Adalimumab (IBI1303) group and 72% of patients in the innovator Adalimumab (Humira™) group [25]. The ASAS 20 index response at week 24 in the current uncontrolled study (intention to treat) was 68% but patients did not receive Bs-ADA beyond 10 weeks in the current study.

A recent AS registry study from India reported significant improvement in pain VAS and BASDAI after 24 weeks use of Bs-ADA (Exemptia™). However, only one third of patients had sufficient data for analysis [26]. None of the patients recorded in the Indian registry reported TB. A total of 124 patients suffering from ulcerative colitis and Crohn’s disease received prolonged treatment with Bs-ADA (Exemptia™) in three Indian studies and nine developed TB [27, 28]. In the current study, 6 patients tested positive for latent TB during the screening process and were excluded (Fig 1). Vigorous use of TB screening seems to have reduced the incident rate of TB in the drug trials of Adalimumab in rheumatic disorders from 1·5 to 0·2 per 100 patient-years [29].

Compared to baseline, the serum TNF α assay showed a progressive reduction, albeit modest, at week 24 and 48; correspondingly IL-17 assay showed a modest rise and then fall (Fig 3). In contrast, IL-6 showed a significant reduction at week 24 (consistent with the CRP assay in Table 3); the assay was much higher at week 48 and probably reflected a rise in CRP along with a conspicuous decline in clinical improvement. Serum assay of pro-inflammatory cytokines was reported in patients of spondyloarthritis treated with anti-TNF agents but is not standard care [30].

Finally, the author (AC) has managed several more patients of AS in clinical practice (CRD) employing short-term use (12-16 weeks) of anti-TNF agents, both innovator, and biosimilar, and has shown clinical improvement similar to the current report without encountering a single case of TB (unpublished). Patients in dire need of the anti-TNF agent but with a positive test for latent tuberculosis are invariably treated with a six-month standard course of INH prophylaxis.

## Conclusion

The current study has demonstrated a remarkable degree of clinical improvement with a 10 week standard regimen (six injections) of Bs-ADA (Exemptia™) in patients suffering from symptomatic active chronic AS with an inadequate response to NSAID. The clinical response persisted for at least 24 weeks in several patients after discontinuation of the Bs-ADA. The regimen was well tolerated and safe. Short-term use of Bs-ADA seems a viable option to treat difficult AS in resource-strained situations such as ours. However, further validation will be required before it can be accepted as a standard treatment modality for AS.

## Data Availability

All data produced in the present work are contained in the manuscript and any further data required will be made available upon reasonable request to the authors 6 months after publication

## Acknowledgment

Arthritis Research Care Foundation-Centre for Rheumatic Diseases CRD), Pune, India, provided generous material and logistic help for the study, and all patients were investigated free of cost as per the protocol. Zydus Cadilla India provided substantial concession to several needy patients in the current study to purchase the Biosimilar Adalimumab (study drug). We are grateful to Mr Sameer Desai, Zydus Cadilla, for providing invaluable support and guidance for this study. Several colleagues (rheumatologist) assisted in the study and namely Dr Abraham Mohan, Dr Kiran Adams and Dr Amit Jain. Mr Sushil Chaudhari provided immense help in administration and data entry. Dr S Sarmukaddam, senior biostatistician, provided expert assistance for the statistical plan and analysis. We remain indebted to our patients who wholeheartedly participated in the study and continue in the ongoing long term follow up.

